# Describing the burden of the COVID-19 pandemic in people with psoriasis: findings from a global cross-sectional study

**DOI:** 10.1101/2021.05.04.21256507

**Authors:** SK Mahil, M Yates, ZZN Yiu, SM Langan, T Tsakok, N Dand, KJ Mason, H McAteer, F Meynell, B Coker, A Vincent, D Urmston, A Vesty, J Kelly, C Lancelot, L Moorhead, H Bachelez, F Capon, CR Contreras, C De La Cruz, P Di Meglio, P Gisondi, D Jullien, J Lambert, L Naldi, S Norton, L Puig, P Spuls, T Torres, RB Warren, H Waweru, J Weinman, MA Brown, JB Galloway, CM Griffiths, JN Barker, CH Smith

## Abstract

**Background:** Indirect excess morbidity has emerged as a major concern in the COVID-19 pandemic. People with psoriasis may be particularly vulnerable to this because of prevalent anxiety and depression, multimorbidity and therapeutic use of immunosuppression.

**Objective:** Characterise the factors associated with worsening psoriasis in the COVID-19 pandemic, using mental health status (anxiety and depression) as the main exposure of interest.

**Methods:** Global cross-sectional study using a primary outcome of self-reported worsening of psoriasis. Individuals with psoriasis completed an online self-report questionnaire (PsoProtect*Me;* Psoriasis Patient Registry for Outcomes, Therapy and Epidemiology of COVID-19 Infection *Me*) between May 2020 and January 2021. Each individual completed a validated screen for anxiety (Generalized Anxiety Disorder-2) and depression (Patient Health Questionnaire-2). Odds ratios (OR) and 95% confidence intervals (CI) were estimated using multivariable logistic regression.

**Results:** 4,043 people with psoriasis (without COVID-19) from 86 countries self-reported to PsoProtect*Me* (mean age 47.2 years [SD 15.1]; mean BMI 27.6kg/m^2^ [SD 6.0], 2,684 [66.4%] female and 3,016 [74.6%] of white European ethnicity). 1,728 (42.7%) participants (1322 [77%] female) reported worsening of their psoriasis in the pandemic. A positive screen for anxiety or depression associated with worsening psoriasis in age and gender adjusted (OR 2.04, 95% CI 1.77-2.36), and fully adjusted (OR 2.01, 95% CI 1.72-2.34) logistic regression models. Female sex, obesity, shielding behaviour and systemic immunosuppressant non-adherence also associated with worsening psoriasis. The commonest reason for non-adherence was concern regarding complications related to COVID-19.

**Conclusions:** These data indicate an association between poor mental health and worsening psoriasis in the pandemic. Access to holistic care including psychological support may mitigate potentially long-lasting effects of the pandemic on health outcomes in psoriasis. The study also highlights an urgent need to address patient concerns about immunosuppressant-related risks, which may be contributing to non-adherence.

## Introduction

The COVID-19 pandemic has led to high mortality and morbidity worldwide. Emerging data highlight an indirect excess morbidity related to public health risk-mitigation efforts such as stay-at-home orders and the re-purposing of healthcare services^1^. Increased risks of mental health disorders and shortfalls in the care of common long-term conditions have been described^2–4^.

Anxiety and depression are highly prevalent in psoriasis and impair quality of life^5,6^. They may also exacerbate cutaneous immune dysregulation in psoriasis through hypothalamic-pituitary-adrenal axis and sympathetic nervous system hyperactivity^7^. Psoriasis clinical guidelines thus recommend routinely screening for anxiety and depression and offering specialist psychological support^8,9^. However, the current reduced access to healthcare services and established increased population mental health burden have led to rising concerns over the physical and psychological sequelae of the pandemic in people with psoriasis.

This study thus sought to better understand the impact of the pandemic on people with psoriasis, to inform immediate priorities for clinical care. We used global self-reported cross-sectional data to characterise the factors associated with worsening psoriasis, with a particular emphasis on the impact of anxiety and depression.

## Methods

### Study design, participants

A cross sectional patient survey (Psoriasis Patient Registry for Outcomes, Therapy and Epidemiology of COVID-19 Infection *Me* [PsoProtect*Me*]^10^), launched on 4th May 2020 and available in 9 languages, was disseminated via social media, patient organisations and clinical networks (Table S1)^11^. The eligibility criterion was any person reporting a clinician-confirmed diagnosis of psoriasis.

### Variables

Variables of interest and questionnaire design were developed and tested by a study group of clinicians, epidemiologists, health data researchers and patient representatives. Questions covered disease severity (whether psoriasis had worsened/improved/remained the same during the pandemic), mental health status (Generalized Anxiety Disorder 2-item scale [GAD-2] and Patient Health Questionnaire-2 [PHQ-2] to screen for anxiety and depression, respectively), adherence (whether, and why, medication was stopped during the pandemic), shielding behaviour (defined as quarantine/staying home/strict distancing in the home/avoiding others), in addition to demographic, socio-economic, and clinical characteristics.

### Statistical methods

Data were extracted on 15th January 2021 and analysed using Stata version 16. After excluding participants self-reporting COVID-19 (a confounding variable), the association between mental health status and worsening psoriasis was assessed using: (a) a minimally adjusted logistic regression model including age and sex covariates; and (b) a fully adjusted model including a consensus list of covariates selected *a priori* as potentially influential on psoriasis severity and anxiety/depression on the basis of expert opinion and existing evidence. In keeping with prior studies, mental health status (anxiety/depression) was defined as a binary variable: participants who scored ≥3 in either GAD-2 or PHQ-2 had a positive screen^12^. Combining GAD-2 and PHQ-2 generates the PHQ-4, a validated measure of mental health status^13,14^. Country of residence was included as a cluster variable in the calculation of standard errors. The fully adjusted model was rerun following multiple imputation using chained equations (20 cycles) to account for missing covariate data. There were no adherence data for those not on systemic therapy, so a separate adjusted model was performed to include adherence.

## Results

A total of 4,043 people with psoriasis from 86 countries (most frequently UK [2,215, 54.8%], USA [243, 6.0%]), Portugal [227, 5.6%] were included. Their mean age was 47.2 years (SD 15.1); mean BMI 27.6kg/m^2^ (SD 6.0), 2,684 (66.4%) were female and 3,016 (74.6%) were of white European ethnicity (Table 1). 1,376 (38.5%) participants had a positive screen for anxiety or depression, of whom 1,052 (76.5%) were female.

**Table 1.** Participant demographics and clinical characteristics stratified by disease state.

SD = standard deviation, BMI = body mass index.
|  | Total | Non-worsening disease | Worsening Disease | p-value |
| --- | --- | --- | --- | --- |
|  | N=4,043 | N=2,315 | N=1,728 |  |
| Shielded | 2,224 (55.1%) | 1,240 (53.8%) | 984 (56.9%) | 0.045 |
| Advised to shield | 742 (33.6%) | 465 (37.5%) | 277 (28.6%) | <0.001 |
| Female gender | 2,684 (66.4%) | 1,362 (59.0%) | 1,322 (76.5%) | <0.001 |
| Age, mean (SD) | 47.2 (15.1) | 49.5 (15.3) | 44.2 (14.3) | <0.001 |
| White European ethnicity | 3,016 (74.6%) | 1,707 (73.7%) | 1,309 (75.8%) | 0.15 |
| BMI, mean (SD) | 27.6 (6.0) | 27.4 (5.8) | 28.0 (6.3) | 0.003 |
| Alcohol >14 units a week | 495 (13.8%) | 295 (15.0%) | 200 (12.3%) | 0.018 |
| Current smoker | 559 (15.8%) | 291 (15.0%) | 268 (16.7%) | 0.18 |
| Full time employed | 1,929 (47.7%) | 1,072 (46.3%) | 857 (49.6%) | 0.038 |
| Household number, mean (SD) | 2.8 (1.8) | 2.8 (1.7) | 2.9 (1.9) | 0.003 |
| Key worker | 1,131 (28.1%) | 595 (25.9%) | 536 (31.1%) | <0.001 |
| Psoriasis severity prior to COVID-19 pandemic |  |  |  | <0.001 |
| Clear | 451 (12.0%) | 299 (14.6%) | 152 (8.9%) |  |
| Nearly clear | 767 (20.4%) | 463 (22.7%) | 304 (17.7%) |  |
| Mild | 989 (26.3%) | 477 (23.3%) | 512 (29.9%) |  |
| Moderate | 892 (23.7%) | 442 (21.6%) | 450 (26.2%) |  |
| Moderate-severe | 480 (12.8%) | 273 (13.4%) | 207 (12.1%) |  |
| Severe | 180 (4.8%) | 90 (4.4%) | 90 (5.2%) |  |
| Systemic therapy |  |  |  | <0.001 |
| No Systemic Therapy | 1,980 (56.2%) | 938 (49.4%) | 1,042 (64.2%) |  |
| Standard Systemic Therapy | 560 (15.9%) | 309 (16.3%) | 251 (15.5%) |  |
| Targeted Therapy | 981 (27.9%) | 652 (34.3%) | 329 (20.3%) |  |
| Non adherent to systemic therapy | 284 (18.5%) | 114 (11.9%) | 170 (29.6%) | <0.001 |
| 1 or more comorbidity | 1,606 (39.7%) | 908 (39.2%) | 698 (40.4%) | 0.45 |
| Anxiety | 1,069 (30.1%) | 408 (21.0%) | 661 (41.0%) | <0.001 |
| Depression | 977 (27.5%) | 392 (20.1%) | 585 (36.5%) | <0.001 |
| Anxiety or Depression | 1,376 (38.5%) | 562 (28.8%) | 814 (50.2%) | <0.001 |

**Table 1.** SD = standard deviation, BMI = body mass index.
|  | **Total** | **Non-worsening disease** | **Worsening Disease** | **p-value** |
| --- | --- | --- | --- | --- |
|  | **N=4,043** | **N=2,315** | **N=1,728** |  |
| **Shielded** | 2,224 (55.1%) | 1,240 (53.8%) | 984 (56.9%) | 0.045 |
| **Advised to shield** | 742 (33.6%) | 465 (37.5%) | 277 (28.6%) | <0.001 |
| **Female gender** | 2,684 (66.4%) | 1,362 (59.0%) | 1,322 (76.5%) | <0.001 |
| **Age, mean (SD)** | 47.2 (15.1) | 49.5 (15.3) | 44.2 (14.3) | <0.001 |
| **White European ethnicity** | 3,016 (74.6%) | 1,707 (73.7%) | 1,309 (75.8%) | 0.15 |
| **BMI, mean (SD)** | 27.6 (6.0) | 27.4 (5.8) | 28.0 (6.3) | 0.003 |
| **Alcohol >14 units a week** | 495 (13.8%) | 295 (15.0%) | 200 (12.3%) | 0.018 |
| **Current smoker** | 559 (15.8%) | 291 (15.0%) | 268 (16.7%) | 0.18 |
| **Full time employed** | 1,929 (47.7%) | 1,072 (46.3%) | 857 (49.6%) | 0.038 |
| **Household number, mean (SD)** | 2.8 (1.8) | 2.8 (1.7) | 2.9 (1.9) | 0.003 |
| **Key worker** | 1,131 (28.1%) | 595 (25.9%) | 536 (31.1%) | <0.001 |
| **Psoriasis severity prior to COVID-19 pandemic** |  |  |  | <0.001 |
| **Clear** | 451 (12.0%) | 299 (14.6%) | 152 (8.9%) |  |
| **Nearly clear** | 767 (20.4%) | 463 (22.7%) | 304 (17.7%) |  |
| **Mild** | 989 (26.3%) | 477 (23.3%) | 512 (29.9%) |  |
| **Moderate** | 892 (23.7%) | 442 (21.6%) | 450 (26.2%) |  |
| **Moderate-severe** | 480 (12.8%) | 273 (13.4%) | 207 (12.1%) |  |
| **Severe** | 180 (4.8%) | 90 (4.4%) | 90 (5.2%) |  |
| **Systemic therapy** |  |  |  | <0.001 |
| **No Systemic Therapy** | 1,980 (56.2%) | 938 (49.4%) | 1,042 (64.2%) |  |
| **Standard Systemic Therapy** | 560 (15.9%) | 309 (16.3%) | 251 (15.5%) |  |
| **Targeted Therapy** | 981 (27.9%) | 652 (34.3%) | 329 (20.3%) |  |
| **Non adherent to systemic therapy** | 284 (18.5%) | 114 (11.9%) | 170 (29.6%) | <0.001 |
| **1 or more comorbidity** | 1,606 (39.7%) | 908 (39.2%) | 698 (40.4%) | 0.45 |
| **Anxiety** | 1,069 (30.1%) | 408 (21.0%) | 661 (41.0%) | <0.001 |
| **Depression** | 977 (27.5%) | 392 (20.1%) | 585 (36.5%) | <0.001 |
| **Anxiety or Depression** | 1,376 (38.5%) | 562 (28.8%) | 814 (50.2%) | <0.001 |

Overall, 1,728 (42.7%) participants reported worsening psoriasis in the pandemic and 314 (7.8%) reported an improvement. A greater proportion of those reporting worsening psoriasis had a positive screen for anxiety or depression (814, 47.1%) compared to those without worsening psoriasis (562 of 1954, 28.8%). The proportion of total participants reporting worsening psoriasis or screening positive for anxiety or depression was stable over time (Fig. 2). A greater proportion of females reported worsening psoriasis (1322 of 2684, 49.3%) compared with males (406 of 1354, 30.0%).

An age and sex-adjusted regression model for worsening psoriasis estimated an odds ratio (OR) of 2.04 (95% CI 1.77-2.36) for those with a positive screen for anxiety or depression, compared to those without a positive screen (Table S2). This association was preserved in the fully adjusted model (OR 2.01, 95% CI 1.72-2.34; Fig. 1, Table S3), and when anxiety and depression were considered separately as primary exposure variables (Tables S4-S5).

**Fig. 1.**
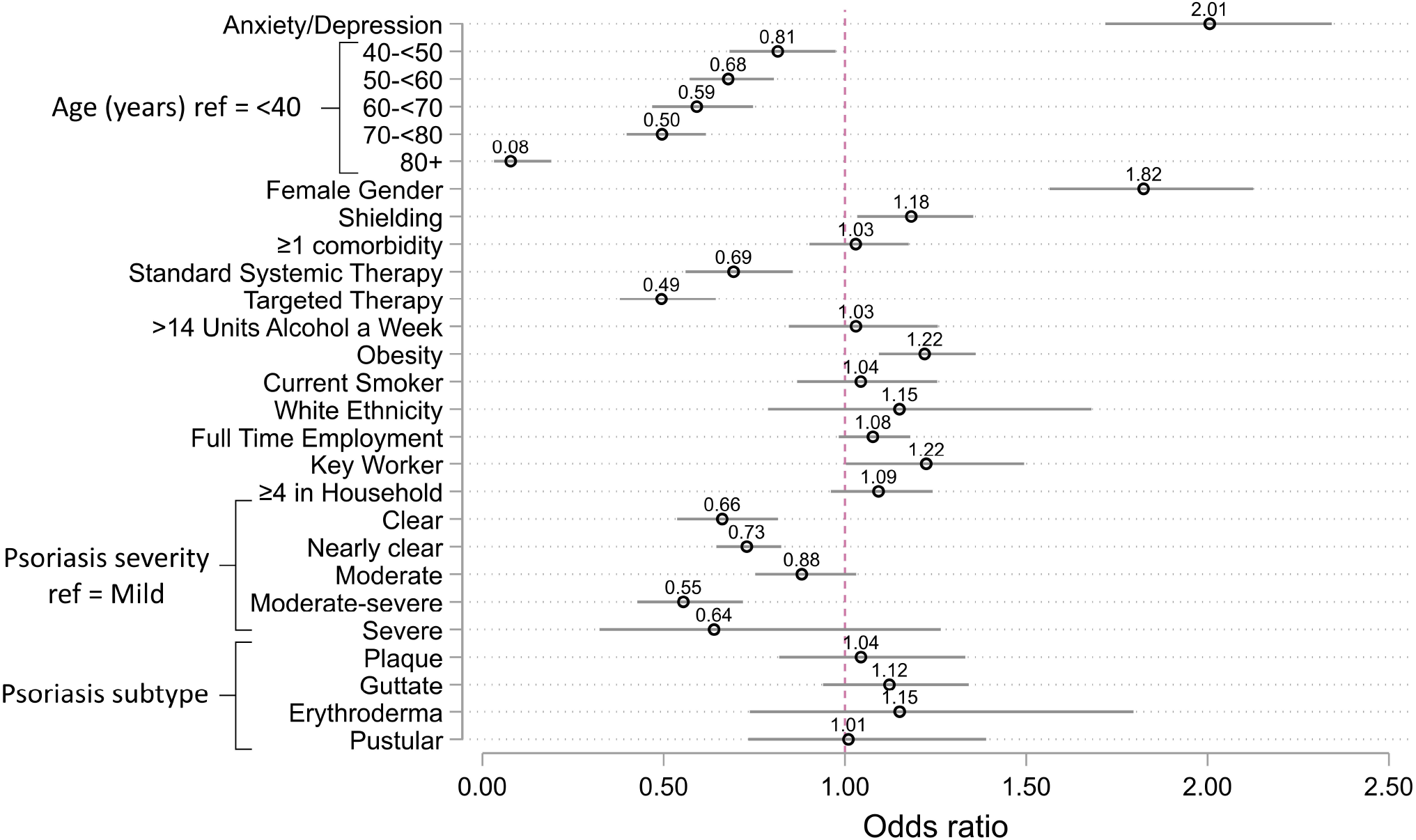
Fully adjusted model for associations with worsening psoriasis. Odds ratios for associations with worsening psoriasis. Anxiety/depression is defined as those who screened positive for either anxiety or depression. Obesity is defined as a BMI greater than 30.

**Fig. 2.**
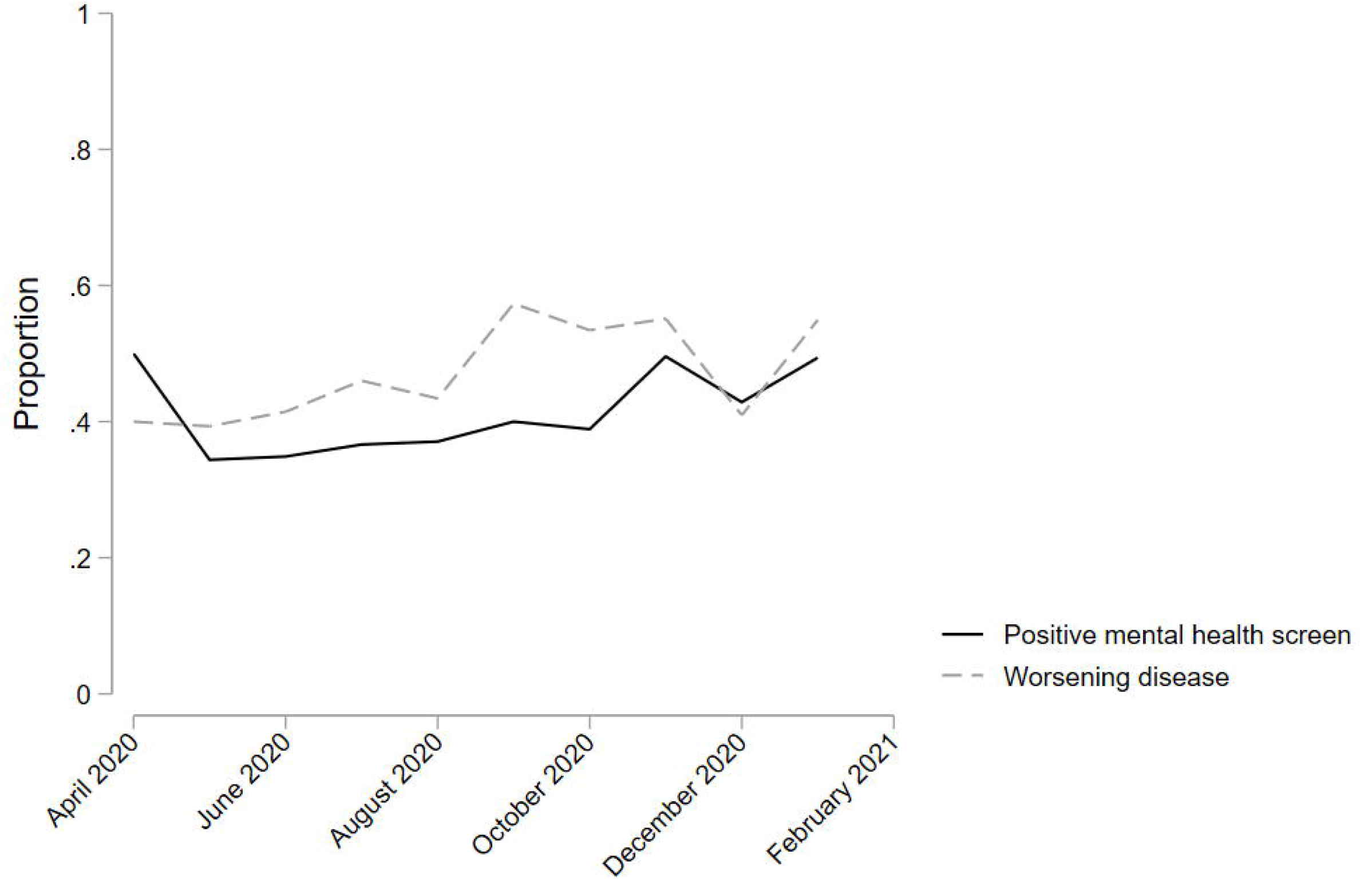
Proportion of respondents with self-reported worsening psoriasis or a positive mental health screen over time

Significant associations with worsening psoriasis in the pandemic were also observed in the fully adjusted model for female gender (OR 1.82, 95% CI 1.56-2.13); obesity (OR 1.22, 95% CI 1.09-1.36); shielding (OR 1.18, 95% CI 1.03-1.35) and key worker status (OR 1.22, 95% CI 1.00-1.49). There were inverse associations with systemic therapy use (standard systemic OR 0.69 [95% CI 0.56-0.86] and targeted therapy OR 0.49 [95% CI 0.38-0.64]).

Of 1,541 (38.1%) participants receiving systemic therapies for their psoriasis, 284 (18.5%) reported non-adherence during the pandemic (Table 1). The commonest reasons for non-adherence were concern regarding complications related to COVID-19 (n=217) and running out of supply (n=48). Therapy non-adherence was associated with worsening psoriasis (OR 2.90, 95% CI 2.31-3.63; Table S6), as estimated using a fully adjusted regression model. A positive screen for anxiety or depression was also more common in those who reported non-adherence compared to those who were adherent (42.8% vs 32.4%).

## Discussion

These self-reported data from >4,000 individuals with psoriasis across 86 countries have immediate clinical relevance. In this population, worsening psoriasis is common and is associated with poor mental health. We also find that in the subset of individuals on systemic therapy, non-adherence is associated with worsening disease and is primarily driven by concerns about immunosuppressant-related risks of COVID-19. This is an important observation since current guidelines generally recommend continuing immunosuppression in people without COVID-19 to maintain disease control^15^. These guidelines are based on reassuring data on drug-related risks of poor COVID-19 outcomes^16^.

While prior studies have investigated the course of COVID-19 and risk factors for severe disease in people with psoriasis^16,17^, there is a paucity of data on patient-reported outcomes in those without COVID-19^18^. Our findings build on data from the general population indicating an increased mental health burden during the pandemic, which is accentuated in females and in those with pre-existing health conditions^19,20^. People with psoriasis may be particularly vulnerable in the pandemic since they have a high prevalence of anxiety and depression compared to the general population, which is further elevated in females and in those with severe psoriasis^21^.

There is evidence to suggest that poor mental health can contribute to worsening of psoriasis, and research into potential underlying inflammatory mechanisms is ongoing^7^. In the context of the pandemic, this association may be further compounded by poor access to healthcare. While causality cannot be inferred from the current study, exploration of the temporal relationship between poor mental health and worsening disease using longitudinal data will be informative (i.e. does anxiety/depression precede worsening psoriasis and/or is it due, in part, to worsening disease).

Our data also highlight gender inequalities in health outcomes in psoriasis. Although male gender is an established risk factor for severe COVID-19, females may be more susceptible to indirect excess morbidity from the pandemic. Further investigation into sex-specific variation in clinical manifestations of mental health disorders, likelihood of reporting worsening psoriasis, differences in baseline severity and responses to systemic therapy is thus warranted^22,23^.

A large international sample of people with psoriasis receiving a broad range of therapies is included, however the generalizability of results is limited by a large proportion being from the UK, female and of white ethnicity. Although validated screening tools for anxiety and depression are used, there is no validation of other self-reported comorbidities, no follow-up information and limited capture of socio-economic variables. Individuals non-adherent to treatment, with low computer literacy, less anxiety or better experiences of the pandemic may be disinclined to participate, which may introduce ascertainment bias.

Taken together, this study indicates a considerable burden due to the COVID-19 pandemic in people with psoriasis. Our data underscore the importance of holistic models of care and indicate a need to provide access to psychological support^8,9^. In those with worsening disease, the possibility of non-adherence to systemic treatment should be explored. Evidence-based communication around medication-related COVID-19 risks and behavioural approaches for supporting adherence may help address potential fears, anxieties and confusion.

Attention given now to address these immediate priorities for clinical care may mitigate a long-lasting detrimental impact of the pandemic on health outcomes in people with psoriasis.

## Data Availability

Data not publicly available.

## Acknowledgements

We are very grateful to the patients who have contributed to PsoProtect*Me*. We would like to acknowledge the professional and patient organizations who supported or promoted PsoProtect*Me* (Table S1). We are grateful for the input of Prof Lars Iversen, Prof Nick Reynolds, Prof Joel Gelfand, Ms Christine Janus and Ms Melissa Sweeney. We would like to acknowledge the following individuals for help with translating the PsoProtect*Me* survey; Dr Haleema Alfailakawi, Dr Wisam Alwan, Dr Rosa Andres Ejarque, Dr Ines Barbosa, Ms Carmen Bugarin Diz, Ms Katarzyna Grys, Dr Mahira Hamdy El Sayed, Mr Tran Hong Truong, Mr Masanori Okuse, Ms Dagmara Samselska, Ms Isabella Tosi, Ms Ya-Hsin Wang. We are also incredibly thankful to Engine Group UK for their generous creative input and website expertise.

## Funding

We acknowledge financial support from the Department of Health via the National Institute for Health Research (NIHR) Biomedical Research Centre based at Guy’s and St Thomas’ NHS Foundation Trust and King’s College London, the NIHR Manchester Biomedical Research Centre and the Psoriasis Association. The views expressed are those of the author(s) and not necessarily those of the NHS, the NIHR, or the Department of Health and Social Care. SKM is funded by a Medical

Research Council (MRC) Clinical Academic Research Partnership award (MR/T02383X/1). ND is funded by Health Data Research UK (MR/S003126/1), which is funded by the UK MRC, Engineering and Physical Sciences Research Council; Economic and Social Research Council; Department of Health & Social Care (England); Chief Scientist Office of the Scottish Government Health and Social Care Directorates; Health and Social Care Research and Development Division (Welsh Government); Public Health Agency (Northern Ireland); British Heart Foundation; and Wellcome Trust. ZZNY is funded by a NIHR Academic Clinical Lectureship through the University of Manchester. CEMG is a NIHR Emeritus Senior Investigator and is funded in part by the MRC (MR/101 1808/1). CEMG and RBW are in part supported by the NIHR Manchester Biomedical Research Centre. SML is supported by a Wellcome senior research fellowship in clinical science (205039/Z/16/Z). SML is also supported by Health Data Research UK (grant no. LOND1), which is funded by the UK MRC, Engineering and Physical Sciences Research Council, Economic and Social Research Council, Department of Health and Social Care (England), Chief Scientist Office of the Scottish Government Health and Social Care Directorates, Health and Social Care Research and Development Division (Welsh Government), Public Health Agency (Northern Ireland), British Heart Foundation and Wellcome Trust.

## Conflict of interest disclosures

Nothing to disclose: Dr Yates, Dr Dand, Prof. Langan, Dr. Norton, Dr. Tsakok, Dr. Yiu, Dr De La Cruz, Dr. Contreras, Ms. Vesty, Ms. Vincent, Mr. Bola Coker, Ms. Meynell, Dr. Lambert, Prof. Brown, Prof. Naldi. Prof. Barker reports grants and personal fees from Abbvie, grants and personal fees from Novartis, grants and personal fees from Lilly, grants and personal fees from J&J, from null, during the conduct of the study.

Prof. Griffiths reports grants and personal fees from AbbVie, grants from Amgen, grants from BMS, grants and personal fees from Janssen, grants from LEO, grants and personal fees from Novartis, grants from Pfizer, grants from Almirall, grants and personal fees from Lilly, grants and personal fees from UCB Pharma, outside the submitted work.

Prof. Jullien reports personal fees and non-financial support from Abbvie, personal fees and non-financial support from Novartis, personal fees and non-financial support from Janssen-Cilag, personal fees and non-financial support from Lilly, personal fees and non-financial support from Leo-Pharma, personal fees and non-financial support from MEDAC, personal fees and non-financial support from Celgene, personal fees from Amgen, outside the submitted work.

Dr. Capon reports consultancy fees from AnaptysBio, grants from Boheringer-Ingelheim, outside the submitted work.

Prof. Bachelez reports personal fees from Abbvie, personal fees from Janssen, personal fees from LEO Pharma, personal fees from Novartis, personal fees from UCB, personal fees from Almirall, personal fees from Biocad, personal fees from Boehringer-Ingelheim, personal fees from Kyowa Kirin, personal fees from Pfizer, outside the submitted work.

Prof. Gisondi reports personal fees from Abbvie, Amgen, Eli Lilly, Janssen, Novartis, Pierre Fabre, Sandoz, UCB, outside the submitted work.

Dr. Galloway reports personal fees from Abbvie, personal fees from Sanofi, personal fees from Novartis, personal fees from Pfizer, grants from Eli Lilly, personal fees from Janssen, personal fees from UCB, outside the submitted work.

Prof. Weinmann has presented talks for Abbvie, Abbott, Bayer, Chiesi, Boehringer Ingelheim, Roche and Merck.

Dr. Mason reports personal fees from LEO Pharma and Novartis, outside the submitted work.

Ms. Moorhead reports personal fees from Abbvie, personal fees from Celgene, personal fees from Janssen, personal fees from LEO Pharma, personal fees from Novartis, personal fees from UCB, outside the submitted work.

Dr. Puig reports grants and personal fees from AbbVie, grants and personal fees from Almirall, grants and personal fees from Amgen, grants and personal fees from Boehringer Ingelheim, personal fees from Bristol Myers Squibb, personal fees from Fresenius-Kabi, grants and personal fees from Janssen, grants and personal fees from Lilly, personal fees from Mylan, grants and personal fees from Novartis, personal fees from Pfizer, personal fees from Sandoz, personal fees from Sanofi, personal fees from Samsung-Bioepis, grants and personal fees from UCB, outside the submitted work.

Dr. Mahil reports departmental income from Abbvie, Celgene, Eli Lilly, Janssen-Cilag, Novartis, Sanofi, UCB, outside the submitted work.

Dr. Di Meglio reports grants and personal fees from UCB, personal fees from Novartis, personal fees from Janssen, outside the submitted work.

Prof. Warren reports grants and personal fees from Abbvie, grants and personal fees from Celgene, grants and personal fees from Eli Lilly, grants and personal fees from Novartis, personal fees from Sanofi, grants and personal fees from UCB|, grants and personal fees from Almirall, grants and personal fees from Amgen, grants and personal fees from Janssen, grants and personal fees from Leo, grants and personal fees from Pfizer, personal fees from Arena, personal fees from Avillion, personal fees from Bristol Myers Squibb, personal fees from Boehringer Ingelheim, outside the submitted work.

Prof. Smith reports grants from Abbvie, grants from Sanofi, grants from Novartis, grants from Pfizer, outside the submitted work.

Dr. Torres reports grants and personal fees from AbbVie, Almirall, Amgen, Arena Pharmaceuticals, Biogen, Biocad, Boehringer Ingelheim, Bristol-Myers Squibb, Celgene, Eli Lilly, Janssen, LEO Pharma, MSD, Novartis, Pfizer, Samsung-Bioepis, Sandoz, during the conduct of the study.

Dr. Waweru is on the Board of the International Federation of Psoriasis Associations who have received grants from Abbvie, Almirall, Amgen, Bristol Meyers Squibb, Boehringer Ingelheim, Celgene, Janssen, Leo Pharma, Eli Lilly, Novartis, Sun Pharma, Pfizer, and UCB, outside the submitted work.

Mr. Urmston reports grants from Almirall, grants from Abbvie, grants from Amgen, grants from Celgene, grants from Dermal Laboratories, grants from Eli Lilly, grants from Janssen, grants from LEO Pharma, grants from T and R Derma, grants from UCB, outside the submitted work.

Ms. McAteer reports grants from Abbvie, grants from Almirall, grants from Amgen, grants from Celgene, grants from Dermal Laboratories, grants from Eli Lilly, grants from Janssen, grants from LEO Pharma, grants from UCB, grants from T and R Derma, outside the submitted work.

## Notes

### Author Declarations

Research approved by KCL research ethics committee (REC ref 20/YH/0135) All necessary patient/participant consent has been obtained and the appropriate institutional forms have been archived.

