## Supplementary file for "Describing the burden of the COVID-19 pandemic in people with psoriasis: findings from a global cross-sectional study"

**Supplementary material**

**Table S1. Organizations who supported or promoted PsoProtect*Me***

| **Organization** |
| --- |
| Psoriasis Association |
| European Society for Dermatological Research (ESDR) |
| International Psoriasis Council (IPC) |
| American Academy of Dermatology (AAD) |
| International Federation of Psoriasis Associations (IFPA) |
| Global Psoriasis Atlas (GPA) |
| Global Skin |
| National Psoriasis Foundation (NPF) |
| European Dermatology Forum (EDF) |
| International League of Dermatological Societies (ILDS) |
| British Association of Dermatologists (BAD) |
| Skin Inflammation and Psoriasis International Network (SPIN) |
| British Society for Investigative Dermatology (BSID) |
| European Academy of Dermatology and Venereology (EADV) |
| Irish Skin Foundation (ISF) |
| Psoriasis and Psoriatic Arthritis Alliance (PAPAA) |
| European Rare and Severe Psoriasis Expert Network (ERASPEN) |
| PSONET |
| British Skin Foundation (BSF) |
| European Umbrella Organisation for Psoriasis Movements (EUROPSO) |
| British Dermatological Nursing Group (BDNG) |
| Australasian Psoriasis Registry (APR) |
| Canadian Psoriasis Network (CPN) |
| French Psoriasis Research Group (Group de Recherche sur le Psoriasis, GRPSO) |
| Canadian Association of Psoriasis Patients (CAPP) |
| Amicus Foundation Psoriasis and PsA (Poland) |
| Civil Association for Psoriasis Patients (Asociación Civil para el Enfermo de Psoriasis, AEPSO, Argentina) |
| Danish Psoriasis Association (Psoriasisforeningen) |
| Finnish Psoriasis Association (Psoriasisliitto) |
| France Psoriasis |
| Fundación de Apoyo a Pacientes con Psoriasis (FUNAPAPSO, Dominican Republic) |
| Global Healthy Living Foundation (GHLF) |
| Hong Kong Psoriasis Patients Association |
| Japan Psoriasis Association (Inspire Japan WPD) |
| Psoriasis Action (Acción Psoriasis) |
| Psoriasis Association of Singapore |
| Psoriasis Group of the Spanish Academy of Dermatology and Venereology |
| Psoriasis New Life Association from El Salvador (Asociacion Psoriasis Nueva Vida El Salvador, PSONUVES) |
| Psoriasis of Panama Foundation (Fundacion Psoriasis de Panama) |
| Psoriasis Philippines (PsorPhil) |
| PsorViet (Vietnam) |
| Psychodermatology UK |
| Puerto Rican Association for Helping Psoriasis Patients (Asociacion Puertorriquena de Ayuda al Paciente de Psoriasis, APAPP) |
| Swedish Psoriasis Association (Psoriasisforbundet) |
| Union of Psoriasis and PsA Associations (Poland) |
| Uruguay Psoriasis Association (Asociación Psoriasis Uruguay, APSUR) |
| Venezuelan Association of Psoriasis (Asociación Venezolana de Psicología Social, AVEPSO) |
| British Association of Dermatologists Biologics Interventions Register (BADBIR) |
| SECURE-AD |
| SECURE-Alopecia |
| SECURE-IBD |

**Table S2. Age and sex-adjusted regression model for worsening psoriasis.**

|  | **Odds Ratio** | **p-value** | **[95% Confidence Interval]** | |
| --- | --- | --- | --- | --- |
| Mental health screen positive | 2.04 | <0.001 | 1.77 | 2.36 |
| Age (years) |  |  |  |  |
| 16-<40 (ref) | 1.00 | . | . | . |
| 40-<50 | 0.80 | 0.02 | 0.66 | 0.96 |
| 50-<60 | 0.68 | <0.001 | 0.57 | 0.83 |
| 60-<69 | 0.53 | <0.001 | 0.43 | 0.67 |
| 70-<79 | 0.48 | <0.001 | 0.35 | 0.65 |
| 80+ | 0.13 | <0.001 | 0.05 | 0.34 |
| Female gender | 1.97 | <0.001 | 1.69 | 2.29 |

**Table S3. Fully adjusted regression model for worsening psoriasis.**

|  | **Odds Ratio** | **p-value** | **[95% Confidence Interval]** | |
| --- | --- | --- | --- | --- |
| Mental health screen positive | 2.01 | <0.001 | 1.72 | 2.34 |
| Age (years) |  |  |  |  |
| 16-<40 (ref) | 1.00 | . | . | . |
| 40-<50 | 0.80 | 0.02 | 0.66 | 0.96 |
| 50-<60 | 0.68 | <0.001 | 0.57 | 0.83 |
| 60-<69 | 0.53 | <0.001 | 0.43 | 0.67 |
| 70-<79 | 0.48 | <0.001 | 0.35 | 0.65 |
| 80+ | 0.13 | <0.001 | 0.05 | 0.34 |
| Female gender | 1.82 | <0.001 | 1.56 | 2.13 |
| Shielding | 1.18 | 0.01 | 1.03 | 1.35 |
| 1 or more comorbidity | 1.03 | 0.67 | 0.90 | 1.18 |
| Systemic treatment |  |  |  |  |
| Nil systemic (ref) | 1.00 | . | . | . |
| Standard systemic | 0.69 | <0.001 | 0.56 | 0.86 |
| Targeted therapy | 0.49 | <0.001 | 0.38 | 0.64 |
| Alcohol >14 units a week | 1.03 | 0.77 | 0.84 | 1.26 |
| Obesity | 1.22 | <0.001 | 1.09 | 1.36 |
| Current smoker | 1.04 | 0.65 | 0.87 | 1.25 |
| White ethnicity | 1.15 | 0.47 | 0.79 | 1.68 |
| Full-time employment | 1.08 | 0.11 | 0.98 | 1.18 |
| Key worker status | 1.22 | 0.05 | 1.00 | 1.49 |
| Household 4 or more | 1.09 | 0.18 | 0.96 | 1.24 |
| Psoriasis severity |  |  |  |  |
| Clear | 0.66 | <0.001 | 0.54 | 0.82 |
| Nearly clear | 0.73 | <0.001 | 0.65 | 0.82 |
| Mild (ref) | 1.00 | . | . | . |
| Moderate | 0.88 | 0.11 | 0.75 | 1.03 |
| Moderate/severe | 0.55 | <0.001 | 0.43 | 0.72 |
| Severe | 0.64 | 0.20 | 0.32 | 1.26 |
| Psoriasis subtype |  |  |  |  |
| Plaque | 1.04 | 0.73 | 0.82 | 1.33 |
| Guttate | 1.12 | 0.20 | 0.94 | 1.34 |
| Erythroderma | 1.15 | 0.54 | 0.74 | 1.80 |
| Pustular | 1.01 | 0.95 | 0.73 | 1.39 |

**Table S3.** Obesity defined anyone with a BMI >30. Targeted therapy was defined as anyone taking TNF inhibitors (adalimumab, certolizumab, etanercept, infliximab), IL-17 inhibitors (ixekizumab, secukinumab, brodalumab), IL-23 inhibitors (guselkumab, risankizumab, ustekinumab) and apremilast. Standard systemic therapy was defined as anyone taking acitretin, ciclosporin, or methotrexate and not taking a targeted therapy.

**Table S4. Fully adjusted regression model for worsening psoriasis with anxiety as primary exposure variable.**

|  | **Odds Ratio** | **p-value** | **[95% Confidence Interval]** | |
| --- | --- | --- | --- | --- |
| Anxiety screen positive | 2.12 | <0.001 | 1.86 | 2.41 |
| Age (years) |  |  |  |  |
| 16-<40 (ref) | 1.00 | . | . | . |
| 40-<50 | 0.81 | 0.02 | 0.68 | 0.97 |
| 50-<60 | 0.68 | <0.001 | 0.57 | 0.81 |
| 60-<69 | 0.59 | <0.001 | 0.47 | 0.73 |
| 70-<79 | 0.48 | <0.001 | 0.39 | 0.59 |
| 80+ | 0.09 | <0.001 | 0.04 | 0.20 |
| Female gender | 1.80 | <0.001 | 1.55 | 2.08 |
| Shielding | 1.19 | 0.01 | 1.05 | 1.36 |
| 1 or more comorbidity | 1.04 | 0.59 | 0.91 | 1.18 |
| Systemic treatment |  |  |  |  |
| Nil systemic (ref) | 1.00 | . | . | . |
| Standard systemic | 0.69 | <0.001 | 0.56 | 0.85 |
| Targeted therapy | 0.48 | <0.001 | 0.37 | 0.62 |
| Alcohol >14 units a week | 1.04 | 0.68 | 0.85 | 1.29 |
| Obesity | 1.24 | <0.001 | 1.11 | 1.39 |
| Current smoker | 1.05 | 0.62 | 0.87 | 1.25 |
| White ethnicity | 1.17 | 0.41 | 0.80 | 1.70 |
| Full-time employment | 1.06 | 0.23 | 0.96 | 1.17 |
| Key worker status | 1.21 | 0.06 | 1.00 | 1.48 |
| Household 4 or more | 1.09 | 0.19 | 0.96 | 1.25 |
| Psoriasis severity |  |  |  |  |
| Clear | 0.66 | <0.001 | 0.54 | 0.81 |
| Nearly clear | 0.75 | <0.001 | 0.66 | 0.84 |
| Mild (ref) | 1.00 | . | . | . |
| Moderate | 0.88 | 0.09 | 0.76 | 1.02 |
| Moderate/severe | 0.56 | <0.001 | 0.44 | 0.72 |
| Severe | 0.62 | 0.16 | 0.33 | 1.20 |
| Psoriasis subtype |  |  |  |  |
| Plaque | 1.07 | 0.58 | 0.83 | 1.39 |
| Guttate | 1.12 | 0.22 | 0.93 | 1.35 |
| Erythroderma | 1.21 | 0.42 | 0.76 | 1.91 |
| Pustular | 1.01 | 0.95 | 0.73 | 1.40 |

**Table S4.** Obesity defined anyone with a BMI >30. Targeted therapy was defined as anyone taking TNF inhibitors (adalimumab, certolizumab, etanercept, infliximab), IL-17 inhibitors (ixekizumab, secukinumab, brodalumab), IL-23 inhibitors (guselkumab, risankizumab, ustekinumab) and apremilast. Standard systemic therapy was defined as anyone taking acitretin, ciclosporin, or methotrexate and not taking a targeted therapy.

**Table S5. Fully adjusted regression model for worsening psoriasis with depression as primary exposure variable.**

|  | **Odds Ratio** | **p-value** | **[95% Confidence Interval]** | |
| --- | --- | --- | --- | --- |
| Depression screen positive | 1.96 | <0.001 | 1.65 | 2.32 |
| Age (years) |  |  |  |  |
| 16-<40 (ref) | 1.00 | . | . | . |
| 40-<50 | 0.80 | 0.01 | 0.68 | 0.95 |
| 50-<60 | 0.66 | <0.001 | 0.56 | 0.77 |
| 60-<69 | 0.56 | <0.001 | 0.43 | 0.73 |
| 70-<79 | 0.47 | <0.001 | 0.39 | 0.58 |
| 80+ | 0.08 | <0.001 | 0.03 | 0.19 |
| Female gender | 1.93 | <0.001 | 1.65 | 2.25 |
| Shielding | 1.21 | 0.01 | 1.05 | 1.39 |
| 1 or more comorbidity | 1.01 | 0.90 | 0.87 | 1.17 |
| Systemic treatment |  |  |  |  |
| Nil systemic (ref) | 1.00 | . | . | . |
| Standard systemic | 0.70 | <0.001 | 0.56 | 0.88 |
| Targeted therapy | 0.50 | <0.001 | 0.38 | 0.67 |
| Alcohol >14 units a week | 1.02 | 0.85 | 0.84 | 1.24 |
| Obesity | 1.21 | <0.001 | 1.09 | 1.35 |
| Current smoker | 1.04 | 0.68 | 0.87 | 1.24 |
| White ethnicity | 1.16 | 0.44 | 0.80 | 1.69 |
| Full-time employment | 1.08 | 0.13 | 0.98 | 1.20 |
| Key worker status | 1.26 | 0.02 | 1.04 | 1.53 |
| Household 4 or more | 1.11 | 0.11 | 0.98 | 1.25 |
| Psoriasis severity |  |  |  |  |
| Clear | 0.64 | <0.001 | 0.53 | 0.78 |
| Nearly clear | 0.72 | <0.001 | 0.63 | 0.83 |
| Mild (ref) | 1.00 | . | . | . |
| Moderate | 0.89 | 0.13 | 0.76 | 1.04 |
| Moderate/severe | 0.54 | <0.001 | 0.42 | 0.70 |
| Severe | 0.62 | 0.14 | 0.32 | 1.18 |
| Psoriasis subtype |  |  |  |  |
| Plaque | 1.08 | 0.52 | 0.85 | 1.38 |
| Guttate | 1.14 | 0.16 | 0.95 | 1.37 |
| Erythroderma | 1.13 | 0.60 | 0.72 | 1.77 |
| Pustular | 0.98 | 0.89 | 0.71 | 1.34 |

**Table S5.** Obesity defined anyone with a BMI >30. Targeted therapy was defined as anyone taking TNF inhibitors (adalimumab, certolizumab, etanercept, infliximab), IL-17 inhibitors (ixekizumab, secukinumab, brodalumab), IL-23 inhibitors (guselkumab, risankizumab, ustekinumab) and apremilast. Standard systemic therapy was defined as anyone taking acitretin, ciclosporin, or methotrexate and not taking a targeted therapy.

**Table S6. Fully adjusted regression model with only those on systemic treatment.**

|  | **Odds Ratio** | **p-value** | **[95% Confidence Interval]** | |
| --- | --- | --- | --- | --- |
| Mental health screen positive | 2.32 | <0.001 | 1.93 | 2.78 |
| Age (years) |  |  |  |  |
| 16-<40 (ref) | 1.00 | . | . | . |
| 40-<50 | 0.68 | <0.001 | 0.55 | 0.84 |
| 50-<60 | 0.61 | <0.001 | 0.50 | 0.75 |
| 60-<69 | 0.45 | <0.001 | 0.34 | 0.59 |
| 70-<79 | 0.32 | <0.001 | 0.22 | 0.48 |
| 80+ | 0.13 | <0.001 | 0.11 | 0.16 |
| Female gender | 1.64 | <0.001 | 1.37 | 1.95 |
| Shielding | 1.22 | 0.05 | 1.00 | 1.48 |
| 1 or more comorbidity | 1.29 | 0.06 | 0.99 | 1.67 |
| Systemic treatment |  |  |  |  |
| Standard systemic (ref) | 1.00 | . | . | . |
| Targeted therapy | 0.81 | 0.03 | 0.66 | 0.98 |
| Non-adherent to treatment | 2.90 | <0.001 | 2.31 | 3.63 |
| Alcohol >14 units a week | 0.79 | 0.10 | 0.60 | 1.04 |
| Obesity | 1.09 | 0.30 | 0.93 | 1.27 |
| Current smoker | 1.05 | 0.69 | 0.82 | 1.34 |
| White ethnicity | 1.40 | 0.11 | 0.92 | 2.13 |
| Full-time employment | 0.90 | 0.37 | 0.72 | 1.13 |
| Key worker status | 1.16 | 0.43 | 0.80 | 1.67 |
| Household 4 or more | 1.46 | <0.001 | 1.15 | 1.85 |
| Psoriasis severity |  |  |  |  |
| Clear | 0.47 | <0.001 | 0.40 | 0.56 |
| Nearly clear | 0.63 | <0.001 | 0.57 | 0.71 |
| Mild (ref) | 1.00 | . | . | . |
| Moderate | 1.03 | 0.81 | 0.78 | 1.37 |
| Moderate/severe | 0.56 | <0.001 | 0.40 | 0.78 |
| Severe | 0.58 | 0.18 | 0.27 | 1.27 |
| Psoriasis subtype |  |  |  |  |
| Plaque | 1.50 | 0.01 | 1.09 | 2.06 |
| Guttate | 1.29 | 0.05 | 1.00 | 1.66 |
| Erythroderma | 1.13 | 0.52 | 0.79 | 1.62 |
| Pustular | 0.80 | 0.24 | 0.55 | 1.16 |

**Table S6.** Obesity defined anyone with a BMI >30. Targeted therapy was defined as anyone taking TNF inhibitors (adalimumab, certolizumab, etanercept, infliximab), IL-17 inhibitors (ixekizumab, secukinumab, brodalumab), IL-23 inhibitors (guselkumab, risankizumab, ustekinumab) and apremilast. Standard systemic therapy was defined as anyone taking acitretin, ciclosporin, or methotrexate and not taking a targeted therapy.
